# Retrospective evaluation of human genetic evidence for clinical trial success using Mendelian randomization and machine learning

**DOI:** 10.64898/2026.02.19.26346536

**Authors:** Charles N. J. Ravarani, Marius Arend, Hannes A. Baukmann, Justin L. Cope, Margaretha R. J. Lamparter, James K. Sullivan, Roman Fudim, Andreas Bender, Anders Mälarstig, Marco F. Schmidt

**Author notes:** These authors contributed equally to this work (Charles N. J. Ravarani, Marius Arend).

## Abstract

Human genetics has become a cornerstone of drug target discovery, yet the value of Mendelian randomization (MR) for predicting clinical success remains uncertain. Here, we systematically evaluated MR across 11,482 target–indication pairs with documented Phase II clinical outcomes to assess its utility for drug development. We find that MR statistical significance alone does not enrich for Phase II success, in contrast to genome-wide association study (GWAS) support, which confers an increase in success probability. However, this apparent limitation reflects the heterogeneous nature of clinical failure and the fact that MR encodes information beyond *P* values. When MR-derived features, including instrument strength and explained variance, are integrated into machine learning models, predictive performance improves substantially. An MR-informed XGBoost classifier identifies target–indication pairs with a 55% overall approval rate, corresponding to a 6.4-fold enrichment over unstratified programs and a 2.8-fold improvement over GWAS-supported targets in Phase II. Notably, this enrichment is achieved without reliance on statistically significant MR results. Our findings demonstrate that MR is most informative when treated as a graded, context-dependent source of causal evidence rather than a binary hypothesis test, and that its integration with machine learning enables scalable, genetics-informed prioritization of drug targets across the clinical pipeline.

## Introduction

The overall probability that a drug candidate progresses successfully through all three clinical trial phases is only around 10%.^1^ Phase II has the lowest success rate of all three clinical trial phases, with only 30%—this is where clinical efficacy is tested for the first time.^2,3^ This highlights a central challenge in drug development: the difficulty of accurately predicting clinical efficacy of a drug candidate.

Demonstrating clinical efficacy is achieved through a prospective, randomized, placebo-controlled trial, which represents the gold standard for establishing causality. Outside of biomedicine, due to time and cost constraints, causality can only be investigated prospectively to a limited extent. As a result, a variety of statistical methods have been developed to estimate causal relationships retrospectively from observational data—collectively known as causal inference.^4^

In parallel, human genetics has had a profound impact on pharmaceutical development: Naturally occurring genetic mutations in humans—often referred to as "experiments of nature"—can provide insights into the likely efficacy and potential on-target toxicity of drugs that modulate the corresponding protein targets. These genetic variations help establish relationships between perturbation of a target and clinical outcomes.^5^ Analyses have demonstrated that drug targets with genetic support—i.e., genes associated with the disease through genome-wide association studies (GWAS)—are significantly more likely to succeed in Phase II trials.^6–8^ This insight is already being implemented in practice by the pharmaceutical industry; in 2021, two-thirds of FDA-approved drugs had some form of human genetic evidence.^9^

Mutations that cause monogenic diseases have been shown to be strong predictors of clinical efficacy.^8^ This is because the causal link between the gene (product) and the disease is typically well understood and firmly established. Applying causal inference to GWAS data in order to validate drug targets and predict efficacy in polygenic diseases thus represents a logical and increasingly adopted strategy in pharmaceutical research.

Mendelian randomization (MR) is a key causal inference method that uses genetic variants as instrumental variables (IVs) to estimate the causal effect of an exposure (e.g., a biomarker or gene target) on a disease outcome.^10,11^ Because genetic variants are randomly allocated at conception according to Mendel’s laws, they are typically independent of the confounding factors that complicate traditional observational studies—assuming the variant affects the outcome only through the exposure of interest. MR is therefore highly valuable for drug development, as it can help determine not just whether a target is relevant, but whether it should be inhibited or activated.^12^

Although the concept of MR dates back to the 1980s,^13^ its practical application has expanded rapidly in recent years due to the availability of extensive biobank datasets and advances in computational infrastructure.

To enable a large-scale, retrospective application of MR to drug development, we leveraged a dataset curated by Minikel *et al.*^8^, which includes clinical trial outcomes for 25,713 drug target–indication pairs (TIPs). We enriched this dataset with GWAS summary statistics (Fig. 1a): 10,207 blood expression and blood protein quantitative loci (e/pQTL) drug-target biomarker studies, resolving 2,204 distinct gene products (Fig. 1b), as exposures and 1,653 disease studies, corresponding to 413 unique indications (Fig. 1c), as outcomes. This allowed us to map at least one exposure or outcome dataset for 18,386 TIPs in the retrospective trial data (Supplementary Table 1 & 2) enabling MR runs for these TIPs. In addition, we created a random negative control sample of 14,889 random combinations of targets and indications featured in the retrospective data with available genetic datasets but no retrospective data on drug trial success.

**Fig. 1.**
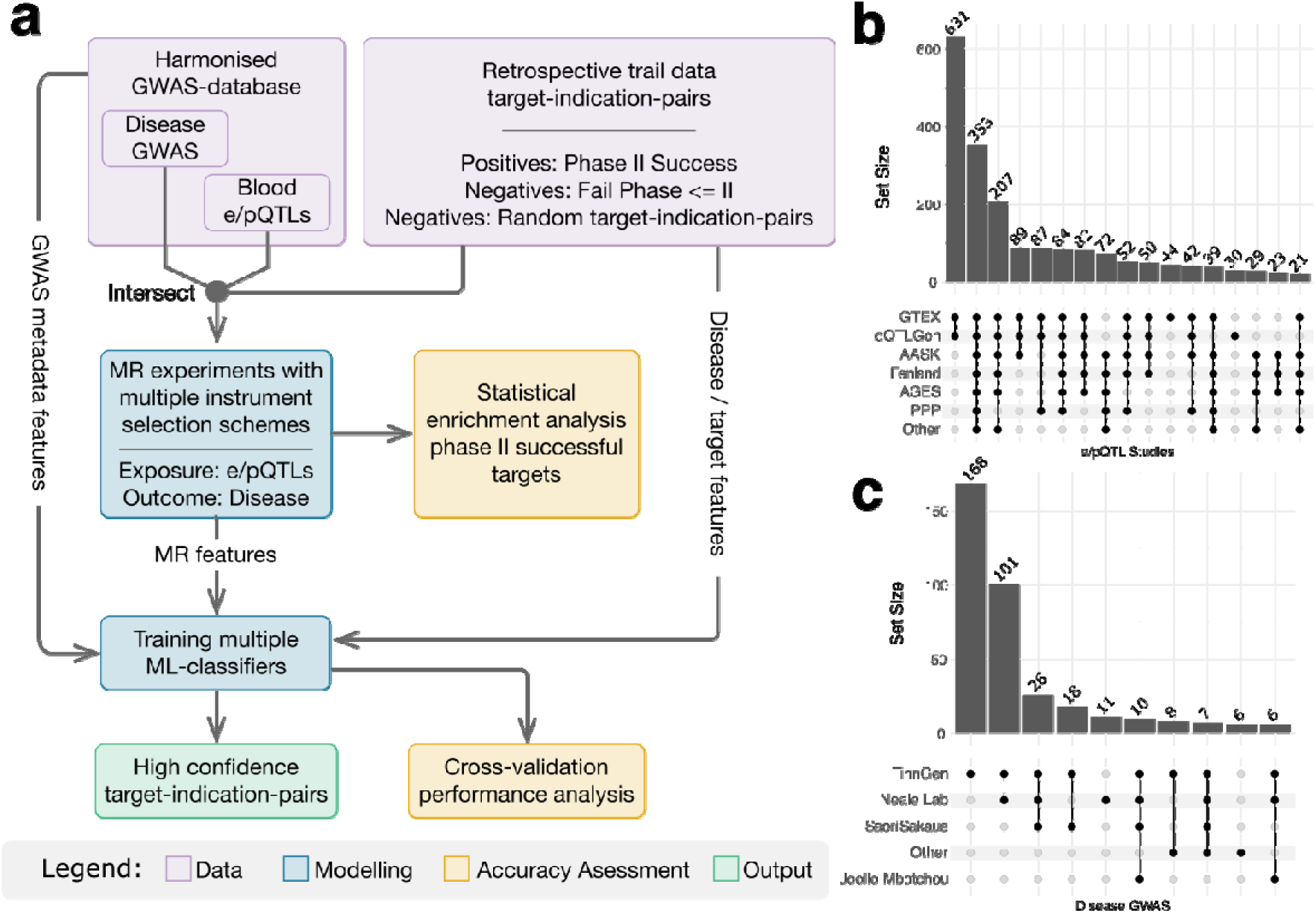
Study design and dataset overview. **(a)** Overview of used data, employed models and validation approaches. **(b)** Upset plot indicating the overlap in coverage for drug-targets resolved in the e/pQTL studies used as exposures (any overlaps including more than 20 targets are shown). **(c)** Upset plot indicating the overlap in coverage for indication resolved in disease GWAS studies used as outcomes (any overlaps including more than 5 indications are shown).

We used our standardized clumping pipeline to investigate MR predictions resulting from instrument sets selected with different clumping parameters (see Methods for details). This dataset formed the basis of our retrospective analysis of the predictive power of MR drug trial outcome prediction. We set out to first check for statistical enrichment of Phase II successful TIPs among significant MR results. Subsequently, we combined these MR results, with additional features from GWAS metadata and drug or disease types and trained different machine learning classifiers to predict Phase II successful TIPs (Fig. 1a). We used a 9-fold crossvalidation scheme to validate our models on unseen Out-Of-Bag (OOB) samples.

Altogether, we present the first large-scale retrospective evaluation of human genetic evidence on clinical trial success using Mendelian randomization and machine learning.

## Results

### Mendelian randomisation analyses of 11,482 target-indication pairs (TIPs)

We identified 18,368 target-indication pairs (TIPs) within the dataset of 25,713 TIPs of successful and failed drug developments curated by Minikel *et al.*^8^, that had both exposure and outcome datasets available. Mendelian randomization analyses were successfully performed for 17,190 TIPs, and 11,482 TIPs had documented outcomes from Phase II clinical trials (Supplementary Table 3), classified as either successful or failed. The results for these 11,482 TIPs are summarized in confusion matrices shown in Fig. 2.

**Fig. 2.**
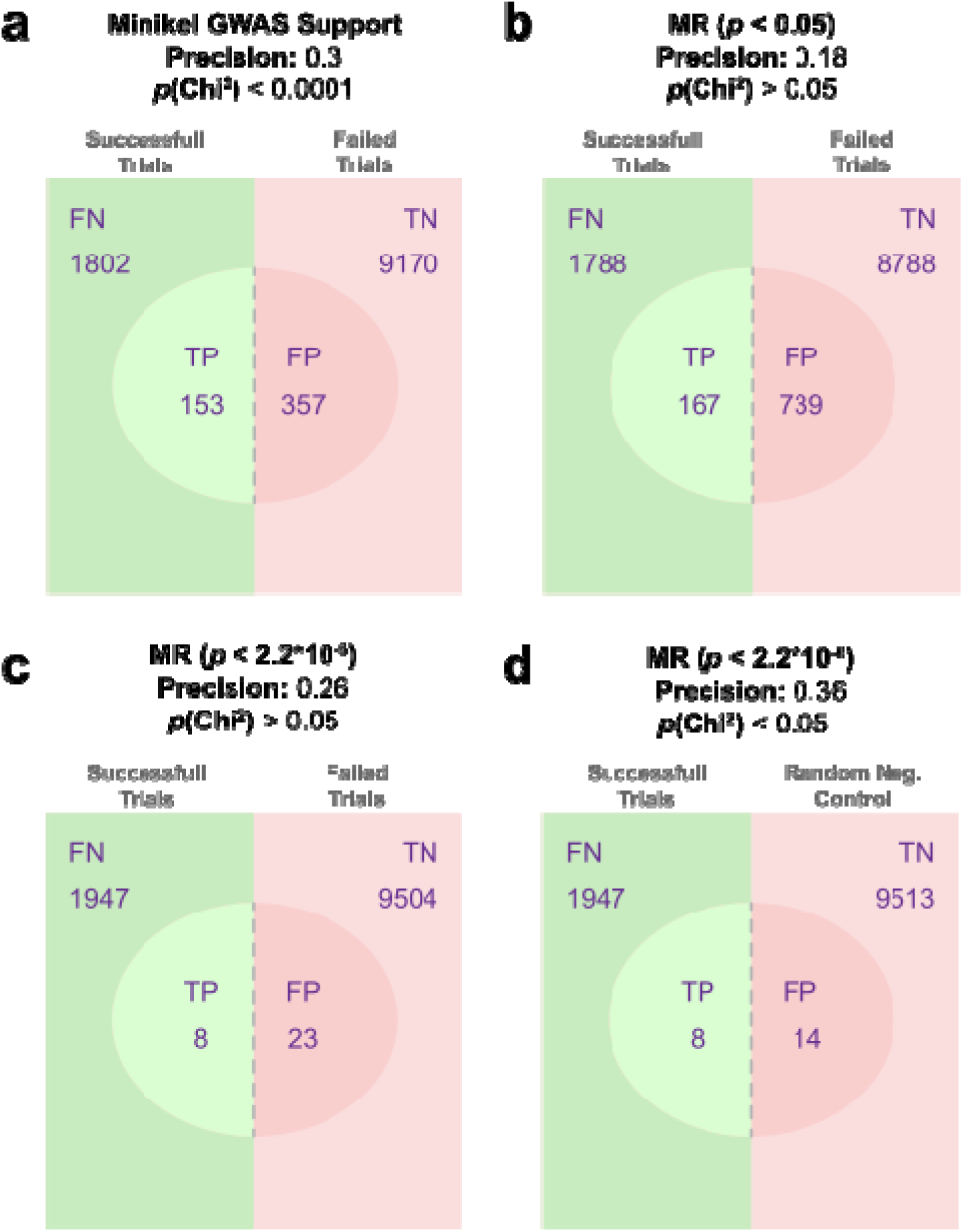
Phase II successful TIPs are enriched in significant MR results and GWAS support. **(a,b)** Confusion matrix and *P* value from chi-squared test results for enrichment of Phase II successful TIPs with **(a)** GWAS support according to Minkel *et al.*^8^ or MR results with below standard significance threshold **(b)** or bonferroni corrected significance threshold. **(c)**. Values are calculated using all TIPs for which retrospective information confirmed failure Prior to Phase III as negatives (Failed trials). **(d)** Confusion matrix and *P* value from chi-squared test results for enrichment of Phase II successful TIPs with significant MR results using a random sample of TIP as negatives. The random sample was filtered for TIPs without retrospective information on Phase II success and without DisGeNET score > 0.3 and stratified for disease area and target type (Random Neg. Control).

TIPs were classified according to the presence or absence of GWAS support, based on the annotation provided by Minikel *et al.*^8^ Among TIPs without GWAS support, the ratio of successful to failed Phase II trials was 1,802:9,170, whereas for TIPs with GWAS support, the ratio was 153:357 (Fig. 2a). This corresponds to a 2.25-fold higher likelihood of Phase II success for TIPs supported by GWAS evidence. The association was highly significant, as determined by a chi-squared test (*P* < 0.0001).

Initial screening with MR using a default significance threshold of *P* < 0.05, resulted in a ratio of 1,788:8,788 successful versus failed trials for TIPs without MR support, compared to 167:739 TIPs with MR support (Fig. 2b). No significant association between MR support and Phase II trial outcome was observed, as indicated by a non-significant chi-squared test.

When applying a Bonferroni-corrected significance threshold (*P* < 2.2 × 10⁻⁶), we did not observe major shifts in the subsequent ratios resulting in 1,947:9,505 for TIPs without MR support and 8:23 for TIPs with MR support (Fig. 2c). Consequently, no significant difference in Phase II success rates was detected based on more stringent MR support to type I errors, as confirmed by a non-significant chi-squared test.

For the amount of compounds that failed primarily due to toxicity or strategic reasons rather than lack of target validity in the retrospective data,^14^ we employed the DisGeNET^15^ gene-disease-association database as a source of biological evidence independent of MR. When mapping DisGeNET scores to the TIPs in the retrospective trial data we could confirm that TIPs bearing a DisGeNET score higher than 0.3 (DisGeNET support) are more successful in clinical trials than the average (Extended Data Fig. 1). Still, the majority of TIPs with DisGeNET support fail prior to Phase II. To generate accuracy estimates that are unaffected by failed compounds with target validity, we constructed a negative set including random combinations of targets and indications from retrospective trial data stratified for the target types and disease areas in Phase II successful TIPs (Supplementary Table 3). Any random combinations with reported Phase II success as well as an associated DisGeNET score higher than 0.3 were filtered out. Using this negative control, the ratio of successful to failed trials was 1,947:9,513 for TIPs without MR support and 8:14 for TIPs with MR support. This resulted in a precision of 0.36 and yielded a statistically significant chi-squared test result, indicating an enrichment beyond random expectation.

### Machine learning using Mendelian randomization–derived features

From the 11,482 TIPs, 10,334 TIPs were used to train two machine learning classifiers, random forest and XGBoost, to distinguish successful from failed Phase II trials, as well as from a random negative control, using 9-fold cross-validation. Features included MR-derived variables (MR method, *P* value, absolute effect size, confidence level, number of instruments, instrument R², and F-statistic), as well as cohort size (exposure and outcome), target class, and disease category (Supplementary Table 3). Precision–recall curves were computed across all out-of-bag (OOB) predictions.

Consistent with expectations, both classifiers showed improved performance when trained against the random negative control compared with failed trials (Fig. 3), with XGBoost consistently outperforming random forest (Extended Data Fig. 2a,b). For the random negative control, XGBoost achieved an AUPR of 0.65 using all features, compared with 0.49 when excluding MR features (Fig. 3a). Similarly, for failed trials, inclusion of MR features improved performance (AUPR 0.46 vs 0.35; Fig. 3b).

**Fig. 3.**
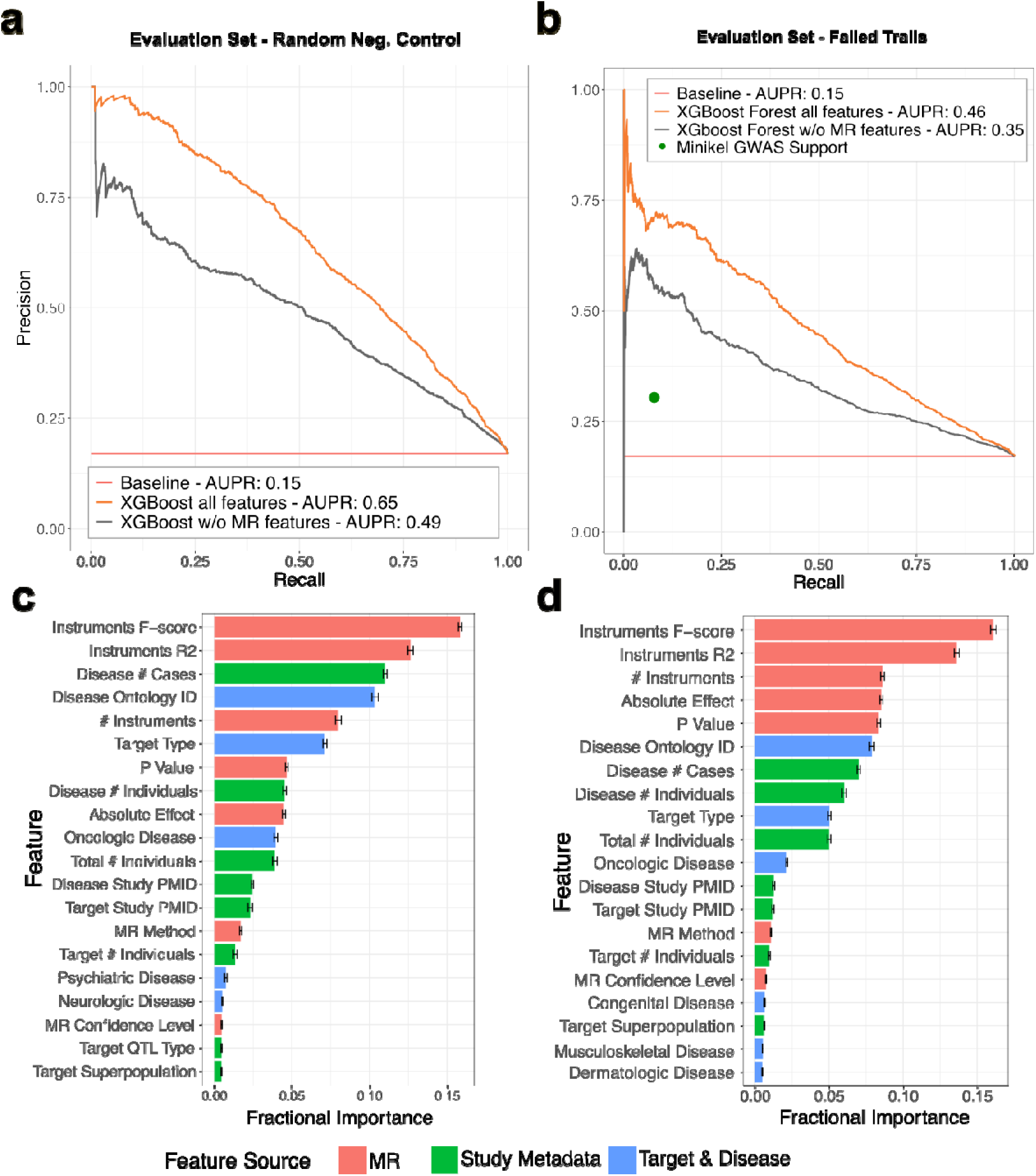
Classification of Phase II successful drugs is improved by Mendelian Randomization features. **(a,b)** Precision recall (PR) curves for the classification of Phase II successful TIPs versus **(a)** randomly generated TIPs for which no retrospective information on Phase II success is available or **(b)** clinically trialed TIPs that failed prior to Phase III. PR curve was calculated over all OOB predictions in 9-fold cross-validation. Curves indicate performance values for a XGBoost classifier using all features (orange), the same model trained without MR features (grey), and the theoretical performance of a classifier that assigns positives randomly based on the number of positives in the validation set (red). The green dot indicates the performance of the binary variable indicating GWAS support from Minikel *et al.*^8^ as a classifier on the same set. **(c,d)** Mean feature importance (fractional gain) from 9-fold cross-validation of XGBoost models trained on the data used in (a) or (b), respectively. Bars give the mean and whiskers the standard error. The color code indicates the source of the feature.

Across both XGBoost models, the most influential features were the instrument F-statistic and instrument R^2^ (Fig. 3c,d). Correlation analysis revealed minimal feature redundancy, with correlation observed only between the number of instruments and the number of individuals in the exposure dataset (Extended Data Fig. 3).

To enable direct comparison, random forest and XGBoost models were additionally trained using GWAS support, as reported by Minikel *et al.*^8^, as a feature. Inclusion of GWAS support did not improve model performance. Overall, MR-derived features consistently provided greater predictive value than GWAS support alone across all machine learning models evaluated (Extended Data Fig. 2a).

### Effect of instrument selection and clumping parameters on classifier performance

To assess the impact of instrument selection and clumping parameters on classifier performance, Mendelian randomization analyses were repeated for all 11,482 TIPs using four additional parameter combinations in addition to the default settings (genome-wide clumping window: 10,000 kb, r² = 0.001, *P* < 5 × 10⁻⁸; Supplementary Table 5). When modulating clumping parameters, we observed differences in classifier performance barely above the standard error from cross validation, independent of the classifier design, negative set or used feature set (Extended Data Fig. 4a,b). Conversely the XGBoost classifier trained on features including MR results depicted the best performance in all four investigated clumping parameter combinations, with the default parameters providing the best basis to segregate successful from Phase II failed TIPs.

When we investigated the association between significant MR results and Phase II successful TIPs (as presented in Fig. 2 for default parameters) a stronger impact of clumping parameters was observed (Extended Data Fig. 5a,b). However, chi-squared tests remain insignificant for all four parameter sets, when segregating failed from Phase II successful retrospective TIPs (Extended Data Fig. 5a).

A different picture emerges when investigating the segregation of random negative TIPs. In this scenario the highly significant chi-squared test result for the default parameter sets is even further improved and the strength of association increases by 31% when reducing the clumping window from the defaull 10,000 kb to 100 (Fig 2d & Extended Data Fig. 5b).

### Retrospective clinical enrichment across genetically and model-prioritized target-indication pairs

Of the 10,334 TIPs used for 9-fold cross-validated machine learning analyses, 9,776 TIPs had available final clinical approval data. As shown in Fig. 4, across all TIPs entering preclinical development, 54% progressed to Phase I (5,315 TIPs), 70% of these advanced to Phase II (3,729 TIPs), 32% progressed to Phase III (1,202 TIPs), and 70% of Phase III programs ultimately achieved regulatory approval (840 TIPs). Overall, 840 of 9,776 TIPs (8.6%) were successful.

**Fig. 4.**
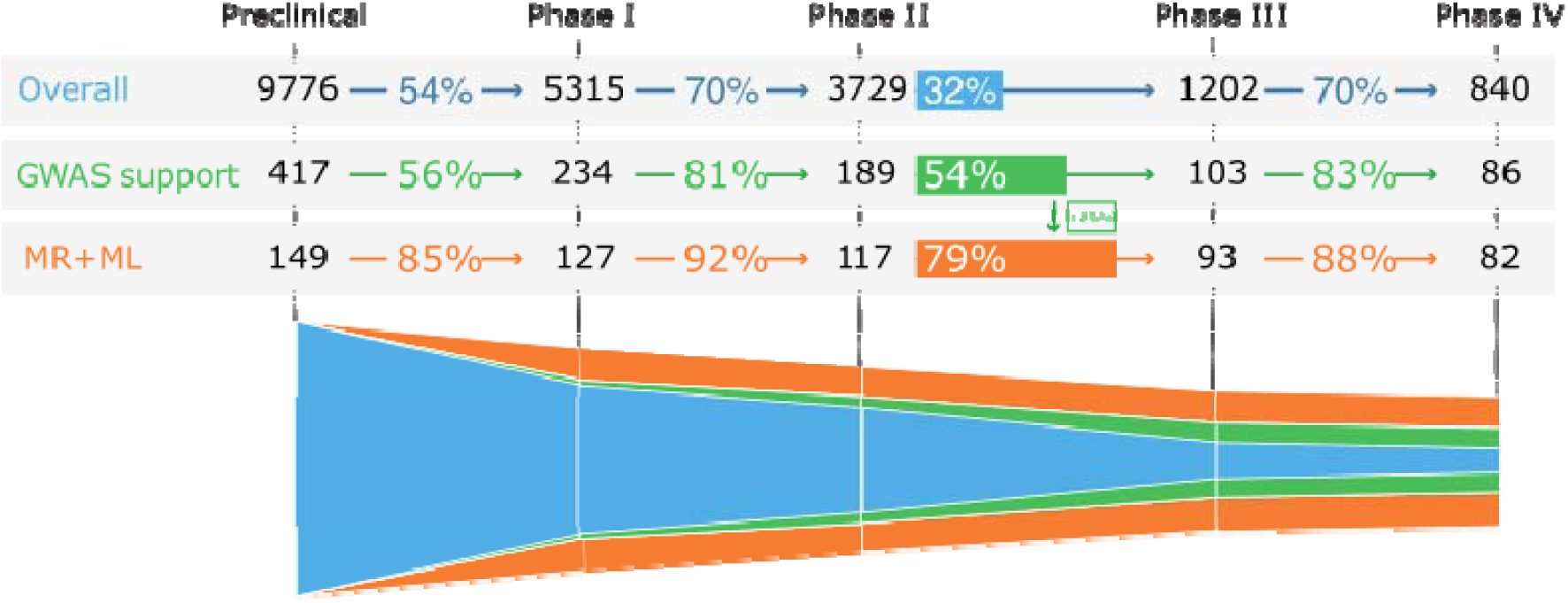
Positives predicted by best MR supported machine learning outperform GWAS supported TIPs. Success chances and funnel chart of different drug sets in the failed trail data set. Overall: all TIPs with retrospective information available (blue, n = 9,776), TIPs with GWAS associated variants according to Minikel *et al.*^8^ (green, n = 417). TIPs predicted as positives in OOB sets by best performing random forest classifier (orange, n = 149). Among 9,776 TIPS, GWAS supported 417 and MR+ML identified 149, with only 12 overlapping TIPs (Jaccard index = 0.02), indicating largely complementary signals.

Among the 417 TIPs with GWAS support (as reported by Minikel *et al.*^8^), progression rates were higher at each stage: 56% reached Phase I (234 TIPs), 81% reached Phase II (189 TIPs), 54% reached Phase III (103 TIPs), and 83% achieved approval (86 TIPs). This corresponds to an overall success rate of 20%, representing a 2.4-fold increase compared with TIPs without GWAS support.

The out-of-bag (OOB) predictions of theXGBoost classifier identified 149 TIPs as successful drug targets. Of these, 85% reached Phase I (127 TIPs), 92% reached Phase II (117 TIPs), 79% reached Phase III (93 TIPs), and 88% achieved approval (82 TIPs), yielding an overall success rate of 55%. Relative to unstratified TIPs, model-supported TIPs were 6.4-fold more likely to succeed and 2.8-fold more likely to succeed than TIPs supported by GWAS alone. Phase II success rates increased from 32% without prior support to 54% with GWAS support and 79% when prioritized by the machine learning model. Notably, among the 9,776 TIPS, 417 were supported by GWAS and 149 were identified by the ML model using MR-informed features, with only 12 overlapping hits; accordingly, the Jaccard index between the GWAS-supported and MR+ML-selected sets was 0.02, indicating that the two approaches capture largely complementary signals.

### Characteristics of top TIPs predicted by the MR-informed classifier

Based on predictions from the best-performing classifier incorporating MR-derived features, 93 Phase II–successful TIPs were identified. The top ten of these TIPs with the highest predicted probability of success are reported in Table 1. Comparison between two most important features contributing to classifier performance and the *P* value from MR analysis showed that none of the predicted successful TIPs were associated with a significant MR result.

**Table 1.** Phase II successful TIPs with highest predicted success probability from best performing classifier. Minikel GWAS Support^8^ is indicated and the associated values of the features, Instrument F-score, Instrument R^2^ and MR *P* value are reported. Rows are ordered by decreasing success probability.

| Rank | Target gene | Indication | Minikel GWAS Support | Instrument R<sup>2</sup> | Instrument F-score | MR *P* value | Success Probability (Classifier) |
| --- | --- | --- | --- | --- | --- | --- | --- |
| 1 | *FLT4* | Gastrointestinal Stromal Tumors | No | 0.084 | 227 | 0.7176 | 0.953 |
| 2 | *TYMS* | Arthritis, Juvenile | No | 0.053 | 37 | 0.6447 | 0.924 |
| 3 | *KIT* | Gastrointestinal Stromal Tumors | Yes | 0.067 | 86 | 0.8569 | 0.921 |
| 4 | *IFNAR2* | COVID-19 | Yes | 0.058 | 491 | 0.1924 | 0.919 |
| 5 | *KIT* | Neuroendocrine Tumors | No | 0.067 | 86 | 0.3470 | 0.915 |
| 6 | *FGFR2* | Carcinoma, Non-Small-Cell Lung | No | 0.015 | 59 | 0.8459 | 0.914 |
| 7 | *F10* | Hemophilia A | Yes | 0.012 | 76 | 0.5354 | 0.912 |
| 8 | *ERBB4* | Carcinoma, Non-Small-Cell Lung | No | 0.049 | 108 | 0.7945 | 0.911 |
| 9 | *IL1B* | COVID-19 | No | 0.018 | 992 | 0.3896 | 0.909 |
| 10 | *TEK* | Carcinoma, Non-Small-Cell Lung | No | 0.112 | 263 | 0.9894 | 0.900 |

The majority of the 93 correctly predicted TIPs comprised kinase targets associated with oncologic indications. In contrast, the five TIPs with the most significant MR results were associated with diverse indications and a variety of targets, each engaged by specific drugs (Supplementary Table 6).

To further characterize these differences, the association between MR significance and Phase II success was evaluated as a function of the number of indications per target. Significant MR results were enriched among successful Phase II targets that had been trialed in a limited number of specific indications (chi-squared test, *P* < 0.001; Extended Data Fig. 6a). This association was not observed for targets that had been trialed in more than five indications. Additionally, targets evaluated across multiple indications were less frequently trialed in metabolic and endocrine diseases and more frequently trialed in dermatologic and oncologic indications (Extended Data Fig. 6b).

## Discussion

In this study, we performed MR analyses across 11,482 TIPs with documented Phase II clinical outcomes to systematically evaluate the utility of genetic causal inference for predicting drug development success. While prior work has demonstrated that GWAS support is associated with increased probability of clinical success, the added value of MR, particularly at scale and across heterogeneous indications, has remained unclear. Our results provide a nuanced answer: MR significance alone does not directly enrich for Phase II success (Fig. 2b,c), yet MR-derived features substantially improve predictive performance when integrated into machine learning models (Fig. 3a,b), yielding strong retrospective clinical enrichment (Fig. 4).

Across the full set of 11,482 Phase II annotated TIPs, we observed no statistically significant association between MR significance and Phase II trial success, regardless of whether nominal (Fig. 2b) or Bonferroni-corrected thresholds (Fig. 2c) were applied. This finding contrasts with the clear enrichment observed for GWAS-supported TIPs, which showed a 2.25-fold increase in Phase II success rates (Fig. 2a). At face value, this result may appear to challenge the utility of MR for target prioritization.

However, several considerations likely explain this apparent discrepancy. First, beyond the MR *P* value, other parameters such as instrument strength (F-statistic), explained variance (R^2^), and consistency across different MR methods carry important information about the reliability and interpretability of genetic evidence.^16^ In this context, direct comparison of MR significance with binary GWAS support using confusion matrices is likely to be misleading. Second, Phase II failure is a heterogeneous endpoint that conflates lack of target validity with non-biological causes such as toxicity, formulation challenges, competitive landscape changes, or strategic portfolio decisions. Consistent with this interpretation, independent biological evidence from DisGeNET demonstrated that many TIPs that fail prior to or during Phase II nevertheless show strong gene–disease associations, suggesting that failure in clinical development does not imply absence of biological relevance (Extended Data Fig. 1). As a consequence, retrospective analyses that treat all failed programs as true negatives are inherently biased against causal inference methods such as MR, which are designed to assess biological validity rather than downstream development feasibility.

Indeed, when we constructed a random negative control explicitly designed to minimize biologically valid target–disease pairs, MR support showed significant enrichment beyond random expectation (Fig. 2d). This result supports the conclusion that MR signals are diluted in conventional retrospective trial datasets by a substantial fraction of biologically valid but clinically unsuccessful programs.

While MR *P* values alone were not predictive of Phase II success, integrating MR-derived features into machine learning models substantially improved performance. Both random forest and XGBoost classifiers consistently benefited from inclusion of MR features, with XGBoost achieving the strongest results (Extended Data Fig. 2a,b). Importantly, MR-derived features outperformed GWAS support in performance (Fig. 3b), and adding GWAS support to the models did not further improve predictive accuracy (Extended Data Figs. 2a, 4a).

The most influential features across all models were the instrument F-statistic and instrument R^2^ (Fig. 3c,d), both of which quantify the strength and explanatory power of the genetic instruments. These features capture information orthogonal to binary GWAS annotations, reflecting not merely the presence of genetic association but the robustness, consistency, and statistical leverage of the underlying genetic architecture. The minimal redundancy observed among numerical features (Extended Data Fig. 4) further indicates that MR-derived variables encode complementary information rather than recapitulating cohort size or target class effects.

These findings suggest that MR should not be viewed as a hypothesis-testing filter at the level of individual TIPs, but rather as a feature generator that encodes graded evidence of genetic causality, which can be exploited by downstream statistical learning approaches.

Systematic evaluation of instrument selection thresholds and linkage disequilibrium clumping parameters demonstrated that classifier performance was generally robust, with default MR settings performing near-optimally (Extended Data Fig. 4a,b). Modest gains were observed for more relaxed *P* value thresholds and reduced gene windows, suggesting that tighter localization of genetic effects may marginally improve signal-to-noise ratios in some contexts. However, these improvements were incremental, indicating that the core conclusions are not sensitive to specific MR parameter choices.

From a practical standpoint, this robustness is encouraging, as it supports the scalability of the approach and reduces the need for extensive parameter tuning when applying MR-informed models in real-world target prioritization settings.

When applied retrospectively across the full drug development pipeline, the MR-informed XGBoost classifier identified a subset of TIPs with dramatically improved clinical outcomes. Model-prioritized TIPs achieved an overall success rate of 55%, representing a 6.4-fold enrichment over unstratified programs and a 2.8-fold enrichment over GWAS-supported TIPs. Phase II success rates increased from 32% in the unstratified set to 54% with GWAS support and to 79% among model-prioritized TIPs (Fig. 4). Across the 9,776 TIPS, GWAS supported 417 and MR+ML identified 149, with only 12 overlapping hits (Jaccard index = 0.02), indicating largely complementary signals. Notably, 11/12 overlapping targets succeeded in Phase II, 9/12 were approved, and only 1/12 stopped in Phase I, suggesting that MR+ML adds orthogonal evidence to GWAS and that concordant hits may be especially strong predictors of clinical success.

These results indicate that integrating MR-derived features with machine learning can identify target–indication pairs that are not only biologically plausible but also disproportionately successful across multiple stages of development, including regulatory approval. Importantly, this enrichment was achieved without relying on statistically significant MR results, underscoring that weak or sub-threshold MR signals can still be highly informative when interpreted in aggregate.

Analysis of the characteristics of top model-predicted TIPs revealed an apparent paradox: the majority of correctly predicted Phase II successful TIPs did not exhibit significant MR *P* values (Table 1). Instead, significant MR results were enriched among targets tested in a limited number of indications (Extended Data Fig. 6), whereas targets trialed broadly across multiple indications, particularly kinases in oncology, rarely showed strong MR significance despite high clinical success rates.

This observation highlights a fundamental limitation of MR in multi-indication settings. When a target is deployed across diverse disease contexts, the genetic signal for any single indication may be diluted or obscured by pleiotropy, context-specific mechanisms, or limited power in individual GWAS datasets. In contrast, MR appears more effective at detecting causal effects in narrow, well-defined indication spaces, where genetic perturbations more closely mirror pharmacologic intervention.

Taken together, these findings suggest that MR significance should be interpreted as context-dependent evidence rather than a universal indicator of target quality, and that its greatest value lies in contributing structured, quantitative features to integrative prioritization frameworks.

This study has several limitations that should be considered when interpreting the results. First, all Mendelian randomization analyses were based on eQTL and pQTL exposure datasets derived from blood. While blood-based molecular QTLs are among the most comprehensive and well-powered resources currently available, gene expression and protein abundance in blood represent a heterogeneous mixture of cell types and may not fully capture tissue- or cell-type–specific regulatory effects relevant to many disease mechanisms. Incorporation of exposure datasets from additional tissues, such as those generated by GTEx^17^ or emerging single-cell QTL resources such as OneK1K^18^, may further refine causal estimates and improve model performance, particularly for indications driven by non-hematopoietic tissues.

Second, we relied on the target–indication annotation curated by Minikel *et al.*^8^, which represents an industry-standard, publicly available dataset and ensures that our analyses are fully reproducible. However, this resource has several important limitations. For a subset of drugs, the true molecular target remains a matter of ongoing scientific debate, leading to potential ambiguity in target assignment. In particular, many small-molecule drugs exhibit polypharmacology and engage multiple targets, a complexity that is not captured by the single target-indication pairs represented in the Minikel dataset. As a result, genetic effects attributed to a single annotated target may, in practice, reflect a composite of on- and off-target mechanisms. In addition to that, this resource does not encode the directionality of pharmacologic modulation, i.e., whether a given drug acts as an inhibitor or activator of its target. As a result, we were unable to leverage one of the key strengths of Mendelian randomization—directional interpretation of causal effects—to align genetic estimates with the mechanism of action of individual compounds. Availability of drug mechanism annotations would likely enhance future models by enabling concordance-based filtering between genetic effect direction and therapeutic modulation, and thereby further improve target prioritization performance. Finally, as with all Mendelian randomization studies, our analyses rely on core assumptions, including the absence of unmeasured horizontal pleiotropy and the validity of linear, lifelong genetic effects as proxies for time-limited pharmacologic intervention, which may not hold uniformly across all targets and indications.

In summary, our results demonstrate that while Mendelian randomization does not, by itself, enrich for Phase II success in retrospective clinical datasets, MR-derived features substantially enhance predictive models of drug development success when combined with machine learning. This approach outperforms GWAS support alone and yields strong enrichment across the full clinical pipeline. These findings argue for a shift in how MR is used in drug discovery: from a binary causal test to a component of multidimensional, data-driven target prioritization strategies and provide a scalable framework for integrating human genetics into translational decision-making.

## Materials & Methods

### Retrospective clinical dataset and genetic annotation

The main source of evidence for drug target success in a certain indication was the information available in the published Supplementary Table 1 of Minikel *et al*.^8^, which indicates the maximum reached phase in completed and ongoing clinical trials for a given target–indication pair (TIP). Detailed information on the original compilation is available per the original publication.^8^ Briefly, Minikel *et al.*^8^ queried the Citeline *Pharmaprojects* database (December 22^nd^ 2022) to retrieve monotherapy programmes added since 2000 annotated with a highest phase reached, a human gene target and a clinical indication defined using the Medical Subject Headings (MeSH) ontology. Combination therapies, programmes without a human target, and entries lacking indication assignments were removed, and each remaining TIP was annotated with the most advanced phase recorded for any drug targeting that gene in that indication. Effectively splitting drugs with multiple targets, into independent target–indication combinations. Minikel *et al.*^8^ generated standardized trial information for a total of 25,713 TIPs.

Minikel *et al.*^8^ also assembled a harmonized database of human genetic associations, aggregating gene–trait links from disparate sources including Online Mendelian Inheritance in Man (OMIM), curated genome-wide association studies (GWAS) and other cohorts, with both indications and genetically associated traits mapped to MeSH terms to standardize phenotypes across datasets. A binary annotation of genetic support was assigned to a TIP if (i) there existed at least one human genetic association linking the target gene to a trait and (ii) the MeSH term for that trait attained a semantic similarity score ≥ 0.8 to the MeSH term for the corresponding clinical indication, as computed via an ontology-based similarity metric. This annotation is referred to as GWAS support when used as a benchmark in this manuscript.

### Disgenet and negative control dataset

As an additional negative evaluation set we generated random combinations of targets and indications included in the retrospective trial data^8^ irrespective of trial outcome. We stratified by target type and disease area distribution in the set of Phase II successful TIPs when drawing random targets and indications, respectively. To filter TIPs with biological evidence from the set of random negative TIPs we sourced information from the DisGeNET database^15^ (March 10^th^ 2022). The DisGeNET database provides a score for ranking of TIPs that summarises evidence from expert curated databases, animal models, clinical trials, human phenotype ontology, and scientific literature mining. We excluded any random combination of target and disease if it was included in the DisGeNET database with a score >0.3 or existed in the set of Phase II successful TIPs.

### Genetic exposure and outcome datasets

To enable MR runs on a scale comparable to the presented evaluation data we compiled a large data set harmonized and standardized GWAS data. We included summary statistics from 10,207 blood e/pQTL datasets resolving 2,204 distinct gene products (Fig. 1b, Supplementary Table 7) and 1,653 disease studies, corresponding to 413 unique indications (Fig. 1c, Supplementary Table 8).

Datasets were pre-processed with version 1.11.8 of the tool MungeSumstats^19^ using default parameters that include checking to ensure variants are on the respective human reference genome (GRCh37 or GRCh38, depending on the dataset), removing duplicate single nucleotide polymorphisms (SNPs), and removing multi-allelic SNPs (according to the SNP database db144).

To facilitate the integration with the TIP evaluation annotation, harmonized and standardized datasets were assigned a Disease Ontology ID for the measured disease indication^20^ or an ENSEMBL Gene ID representing the measured gene product (in the case of eQTL and pQTL datasets). These standardized ids were then used to map datasets to retrospective trial information. To this end, mesh ids in Supplementary Table 1 of Minikel *et al.*^8^ were mapped to Disease Ontology ids using the Disease Ontology and DisGeNET cross-reference database. Gene Names in Supplementary Table 1 of Minikel *et al.*^8^ were mapped to ENSEMBL gene ids using the uniprot API. The mapping allowed to assign each retrospective trial TIP any available e/pQTL datasets for use as exposure and any available disease GWAS datasets for use as outcome in a mendelian randomization experiment, to test for causal effect.

### Mendelian randomization framework

Different clumping parameters were used depending on the scope of the exposure study (cis *vs.* trans QTLs). No other data set-dependent distinctions in clumping parameters were made in order to ensure comparability of the selected instruments between different exposure QTL studies. If not mentioned otherwise the values indicated as “default” in Supplementary Table 5 were used in the subsequently described steps. First, pre-processed summary statistics files were read and thresholds were applied to the exposure SNPs to ensure they were valid instruments. Independent SNPs were identified by performing linkage disequilibrium (LD) clumping to discard SNPs in LD with another variant with a smaller *P* value association based on the European reference panel from the 1,000 Genomes Project^21^ using the ld_clump() function of the R package ieugwasr^22^(version 1.0.1), which provides a wrapper around PLINK^23^. The correlation LD clumping parameter r^2^ and haplotype block size are indicated in the respective results. SNPs remaining after clumping and *P* value thresholding were overlapped with the SNPs reported in the outcome dataset and effect directions were harmonized using the TwoSampleMR package (version 0.6.9)^24^, dropping any palindromic SNPs. The remaining overlapping SNPs represent the list of Instrumental Variables employed in subsequent MR analysis.

Based on the selected *m* Instrumental Variables, R^2^ and F-statistic as measures of their jointly explained variance have been calculated^25^:

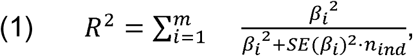

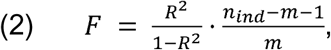

where *β* gives the reported effect size and *n_ind_* the number of individuals in the exposure data set. *SE*(*β*) refers to the standard error of *β*.

If at least three SNPs remained as instrumental variables, the MR-Rücker framework^24^ was applied to determine which random effects modelling framework of the inverse-variance weighted (IVW)^26^ or the MR-Egger^27^ method was best supported by the data and results from the respective MR method were used. If only two SNPs remained as instrumental variables, the IVW method was applied, or if only one, the Wald ratio method^28^. The used method is indicated in Supplementary Table 3 and Supplementary Table 9 gives the distribution of used methods in the results evaluated in this study.All methods were implemented in the TwoSampleMR package^24^ (version 0.6.9).

### Enrichment and validation analyses

In order to evaluate performance and train machine learning classifiers we assigned the TIPs into sets with clear binary annotation indicating whether a TIP was Phase II successful (positive) or not (negative).

The first set is purely based on TIPs with retrospective information on clinical trials. TIPs have been assigned as positives and negatives according to the following rules:

1. TIPs with completed trial status and a maximum reached phase below phase II were assigned as negatives and TIPs with a higher phase as positives.
2. TIPs with ongoing trial status in the database provided by Minikel *et al.*^8^ (queried December 22^nd^ 2022) and a maximum reached phase below phase II were treated as unclassifiable and excluded from the set and TIPs with a higher phase as positives.

The second set combines phase II successful TIPs from retrospective data with a sample of random TIPs as negatives. TIPs in this set have been assigned based on the following rules:

1. TIPs with a maximum reached phase above phase II were assigned as positives. All other TIPs with retrospective trial information were excluded from the set.
2. All randomly sampled TIPs (filtered for DisGeNET score < 0.3) were treated as negatives.

We subsampled a number of random TIPs with MR results that ensured this second evaluation data set has the same cardinality and balance as the first set.

We calculated 2×2 confusion matrices between the evaluation annotation of a TIP (Phase II successful vs negative) and different binary predictors: (i) GWAS support as reported by Minikel *et al.*^8^ or (ii) MR significance at different significance levels *α*. We used established values of 0.05 and 0.0001 as well as, 2.112 x 10^-6^, a Bonferroni corrected value taking into account the number of all MR experiments (The respectively used *α* is indicated in the result figure or the text section). After obtaining the respective confusion matrix, the chi-squared test^29^ was used to test for a significant association between prediction and ground truth. Association strength has been measured using Cramer’s V.

### Machine learning models and feature engineering

To improve on the performance of the raw *P* value as sole predictor of TIP success we trained machine learning classifiers employing different combinations of features from (i) MR experiments, (ii) the metadata of used GWAS studies, (iii) genetic support according to Minikel *et al.*^8^ as well as (iiii) target and disease type (Supplementary Table 4).

As a first step in the creation of training data we split off 10% of the TIPs in the two evaluation data sets as leave-out validation data. The remaining 90% of data were termed training data and used for model training. For hyperparameter tuning and performance analysis the training data was used in a 9-fold cross validation approach.

All machine learning models were trained and evaluated using the R tidymodels framework (version 1.3.0) together with the ranger package (version 0.18.0) for the generation of random forest classifiers^30^ and the XGBoost package (version 3.1.3.1) for the generation of XGBoost classifiers^31^. Hyperparameters were tuned for using the mr results obtained with default clumping parameter and models harbouring metadata and mr features. Hyperparameter tuning was performed employing a 9-fold cross validation approach on 90% of the total data. For the random forest classifiers a grid of values for the hyperparameters number of trees, number of features selected per split and minimum node size were scanned (Supplementary Table 10). For XGBoost classifiers we uniformly sampled 60 hyperparameter combinations from the space spanned by the hyperparameters number of trees, number of features selected per split, minimum node size, learning rate, and minimum loss reduction (Supplementary Table 11). For both classifiers the hyperparameter set maximising the mean AUPRC from crossvalidation was used to train all other machine learning models of the same classifier type.

### Model evaluation and sensitivity analyses

To evaluate the model performance we calculated AUPRC values for each fold and report the mean and standard error as over all cross-validation folds. IPR curves were plotted over the aggregated OOB predictions for each data point in the training data set.

Feature importances presented for XGBoost classifiers in this paper have been calculated as fractional gain,^31^ the relative reduction of variance summed over all splits in which the respective variable was used and weighted over all trees, normalized to the total reduction of variance achieved by the classifier. The fractional gain was calculated using the implementation of the XGBoost R package (version 3.1.3.1). In case of categorical features that have been transformed using one-hot encoding, the feature importances of all resulting features have been summed and are reported as per the original categorical feature. These feature importance calculations have been run for each fold of the cross-validation and mean and standard error over all folds are reported.

### Retrospective clinical progression analysis

To calculate funnel width representing the number of TIPs that were evaluated up to a certain Phase of clinical development we only considered completed trial TIPs in the 90% training data, excluding any annotation from ongoing trials at the time of the citeline database query (December 22^nd^ 2022). Starting from the total set of trialed TIPs in preclinical evaluation we calculate the relative and absolute numbers of these TIPs that were evaluated at each successive stage. We did this for three sets: (a) all TIPs with retrospective information in the training data. (b) The subset of these TIPs that were deemed “genetically supported” in the study from Minikel *et al.*^8^ and (c) another subset of all TIPs with retrospective information that were predicted as successful by the best performing random forest classifier. We choose a probability threshold for success prediction that delivers the same recall as achieved by set (b). If *N*(*i*) indicates the number of drugs evaluated in stage i of the evaluation pipeline we calculated the probability of a random drug in the selection to advance to the next stage, *P*(*i*, *i* + 1), as:

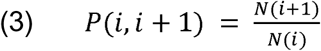

## Data Availability

All data produced in the present study are available upon reasonable request to the authors.

## Acknowledgements

This work was partially supported by Germany’s Federal Agency for Breakthrough Innovation (SPRIN-D) through a validation project. The analyses were supported by NVIDIA Corporation through the *NVIDIA Inception program* and by Google LLC through the *Google for Startups Cloud Program*. We thank Radi Hilaneh for preparing Figures 1–4, and Jack Scannell and Hubert Truebel for proofreading the manuscript.

## Extended Data

**Extended Data Fig. 1.**
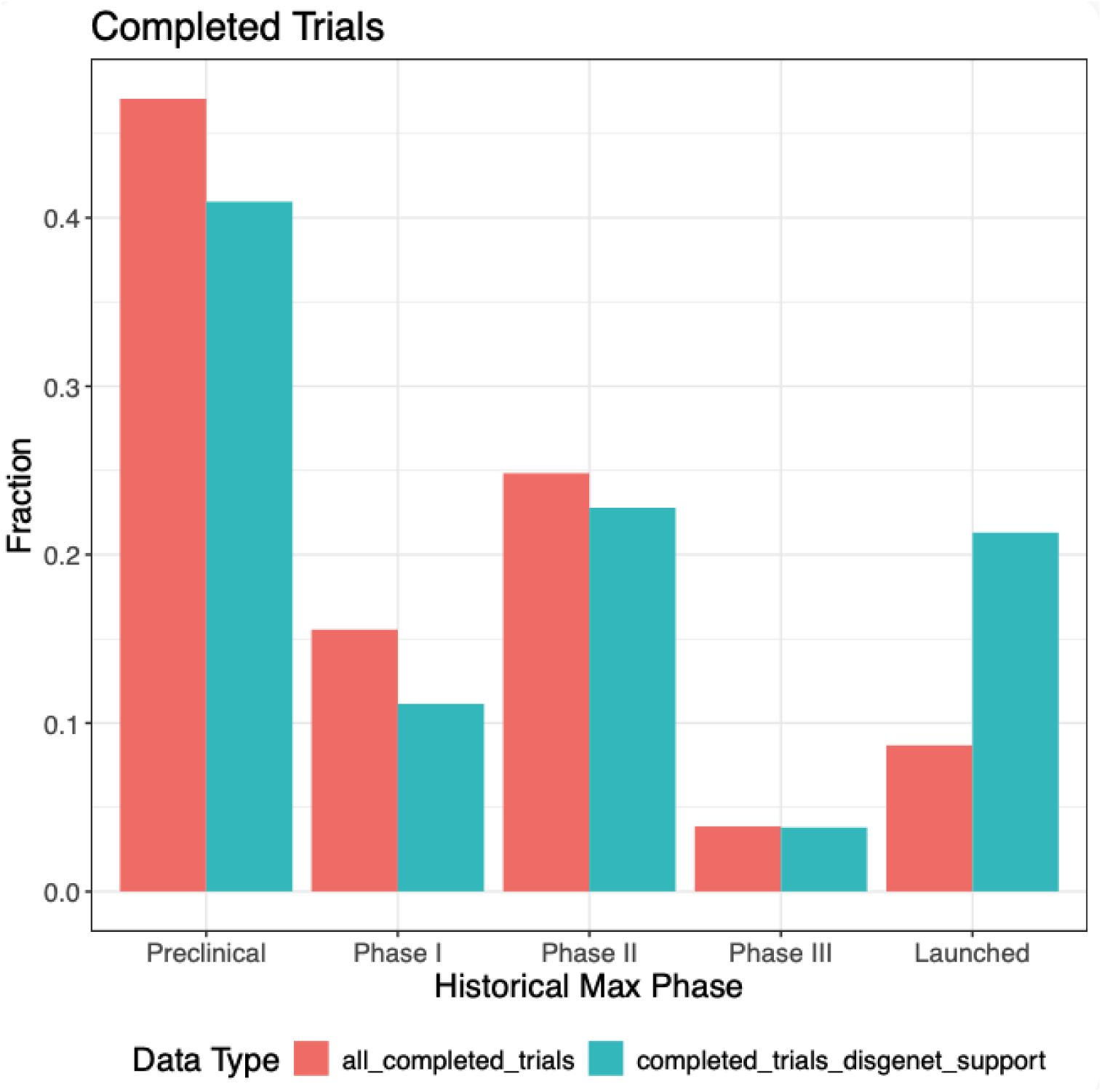
Retrospective trials with DisGeNET support showed a higher proportion of successful TIPs. The barplot visualized the distribution of maximum reached phases for all completed trials in the retrospective data (turquoise) and the subset with DisGeNET support^15^. DisGeNET support was assigned if a TIP is associated with a DisGeNET score higher than 0.3.

**Extended Data Fig. 2.**
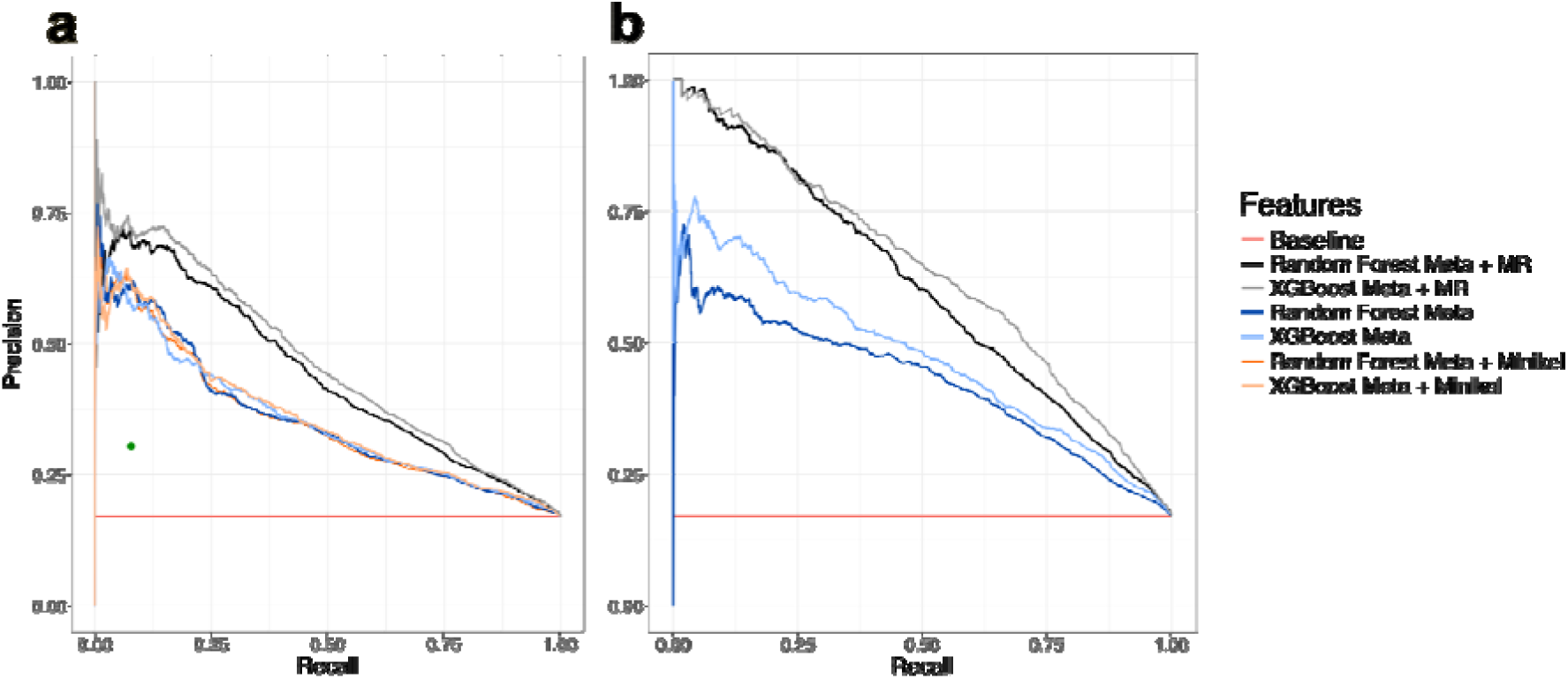
Random Forest and XGBoost classifier depict similar dependency on MR features. **(a)** Precision recall (PR) curves for the classification of Phase II successful drug-target indication pairs (TIPs) versus TIPs that failed prior to Phase III. PR curve was calculated over all Out-Of-Bag prediction in 9-fold cross-validation. Curves indicate performance values for a random forest or XGBoost using different combinations of metadata (Meta), MR result (MR) and Minikel *et al.*^8^ GWAS support features (Minikel). The baseline indicates the theoretical performance of a classifier that assigns positives randomly based on the number of positives in the validation set (baseline). The green dot indicates the performance of the binary variable indicating GWAS support from Minikel et al. as a classifier on the same set. **(b)** Same as in (a) but the negative set consists of random combinations of targets and indications for which no retrospective information on Phase II Success is available, stratified for disease area and target type in the set of Phase II successful TIPs.

**Extended Data Fig. 3.**
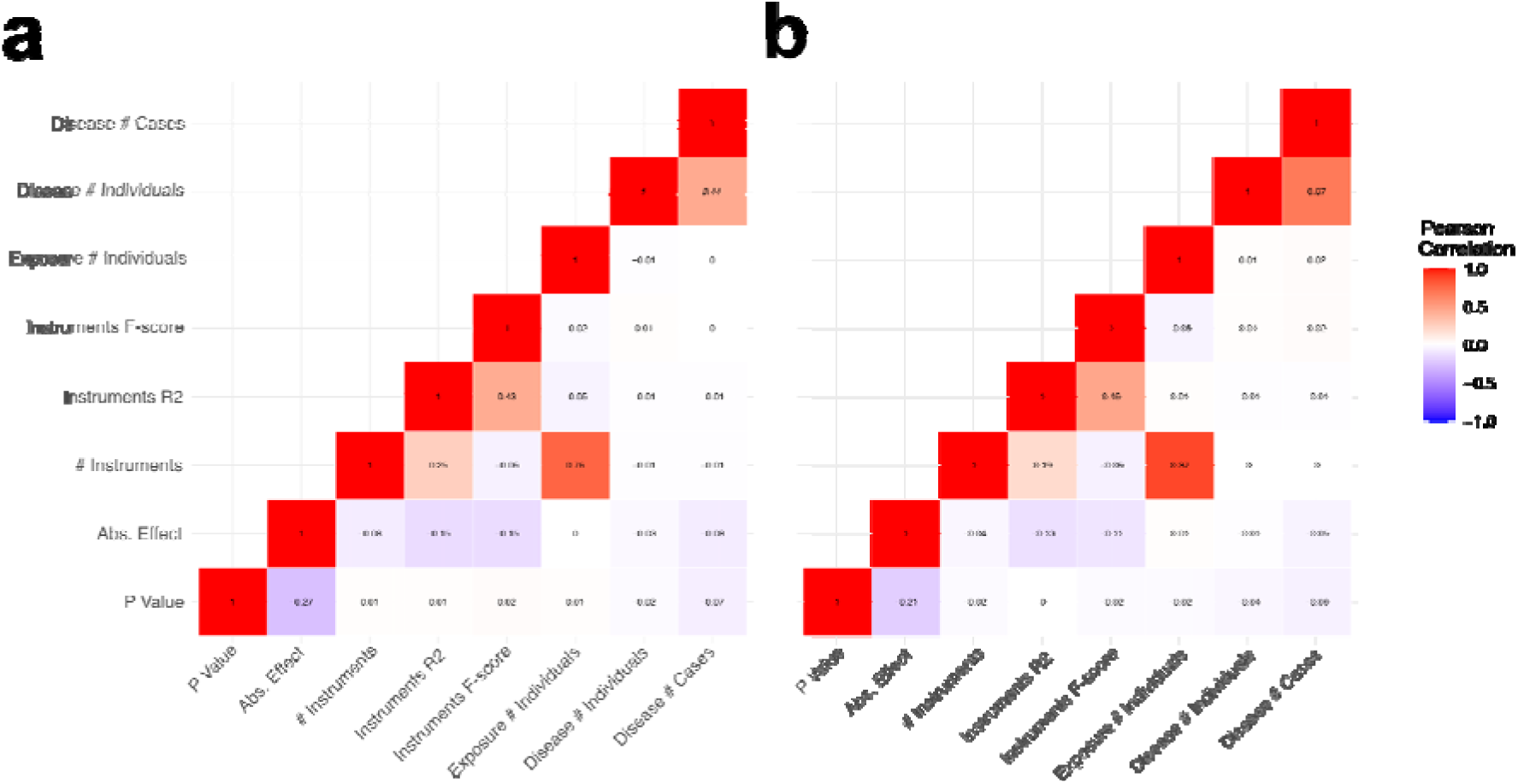
Numerical MR and GWAS derived features show overall low correlation. Correlation matrix of the numerical features derived from MR results and GWAS metadata calculated over the entire training data obtained using the **(a)** random control or **(b)** failed trials as negatives.

**Extended Data Fig. 4.**
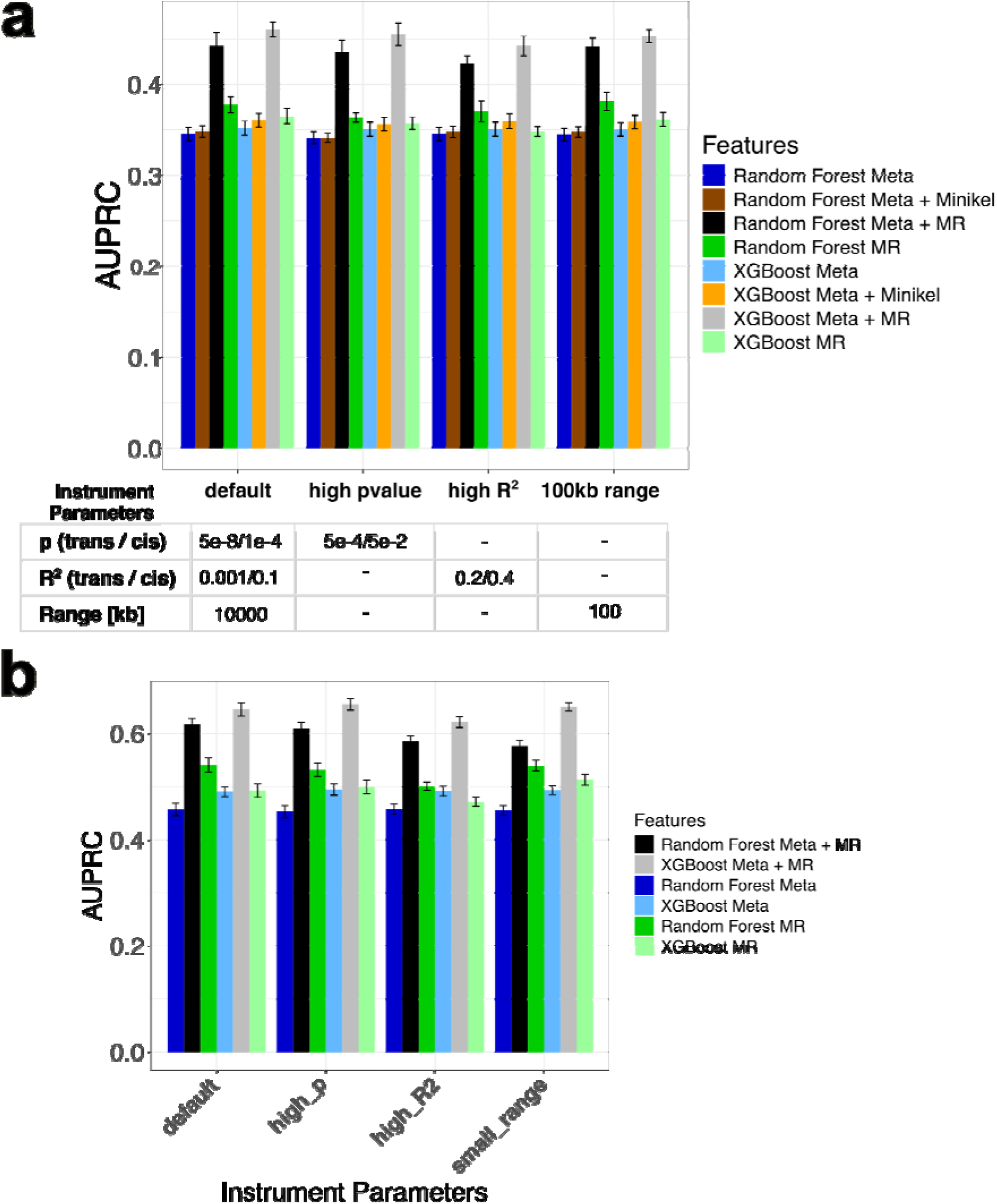
Overview of classifier performance for different models, features and instrument selection schemes. **(a,b)** AUPRC curves for the classification of Phase II successful TIPs versus **(a)** TIPs of failed trials **(b)** random TIPs or **(c)**. AUPR was calculated separately for each OOB set in 9-fold cross-validation of the 90% training data. Bar plot indicates mean and standard error of AUPRC of a random forest or XGBoost classifier using the indicated features (see also **Supplementary Table 4**) The x-axis indicates the different instrument selection scheme used for MR analysis, with the table indicating exact values used for clumping and instrument selection. All models were trained on the intersection of TIPs with available MR results in all instrument selection schemes.

**Extended Data Fig. 5:**
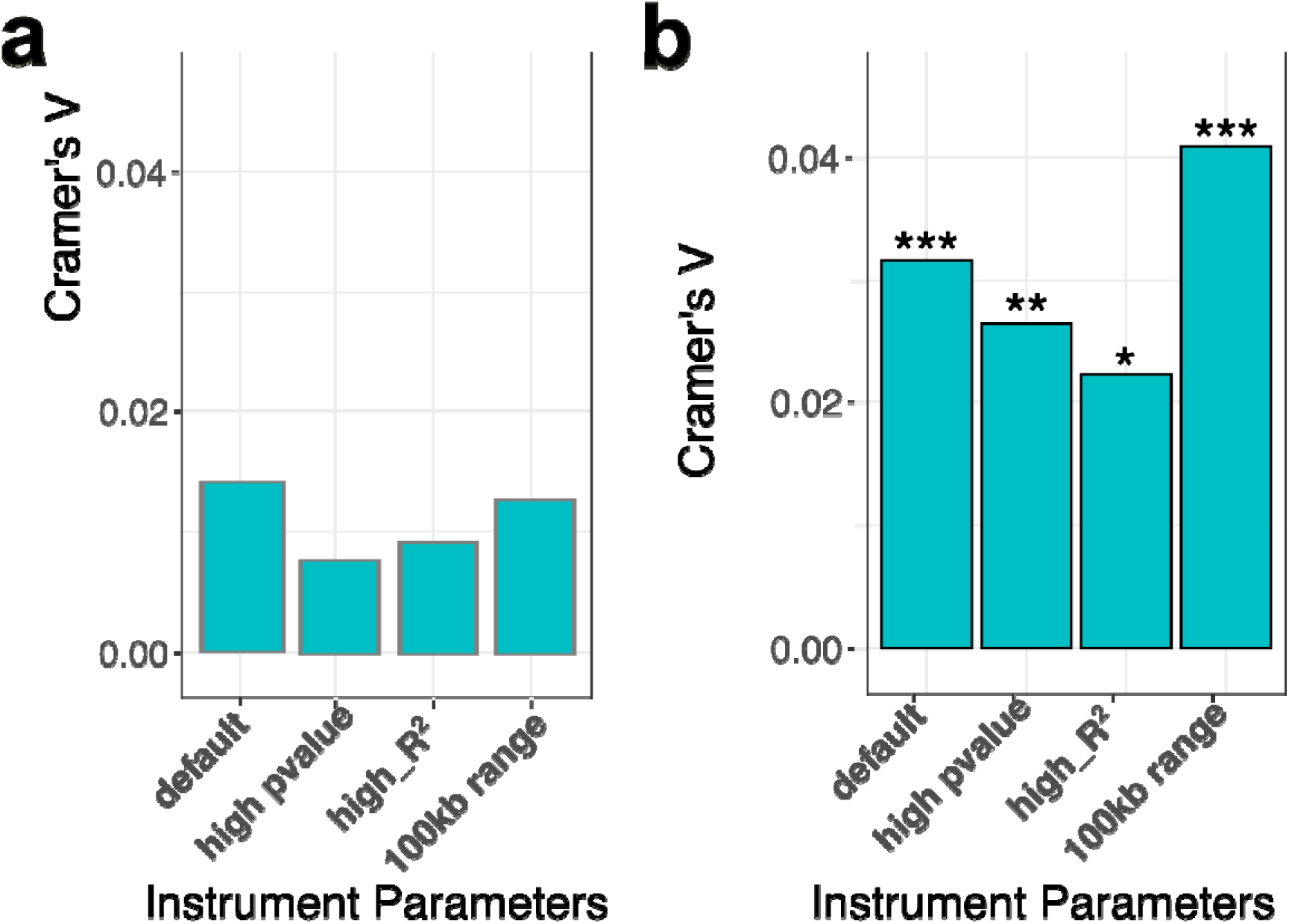
Clumping parameter selection has strong influence on MR significance predictiveness. **(b,d)** Predictive power of the MR *P* value given by Cramer’s V association strength between significant MR results (ɑ = 0.0001) and Phase II successful TIPs vs **(a)** TIPs of failed trials **(b)** random TIPs in the evaluation data. Asterisks indicate associated Chi-squared test significance level. All statistics were calculated on the intersection of TIPs with available MR results in all instrument selection. Significancy asterisk code: * *P* < 0.05; ** *P* <0.01; *** *P* <0.001.

**Extended Data Fig. 6.**
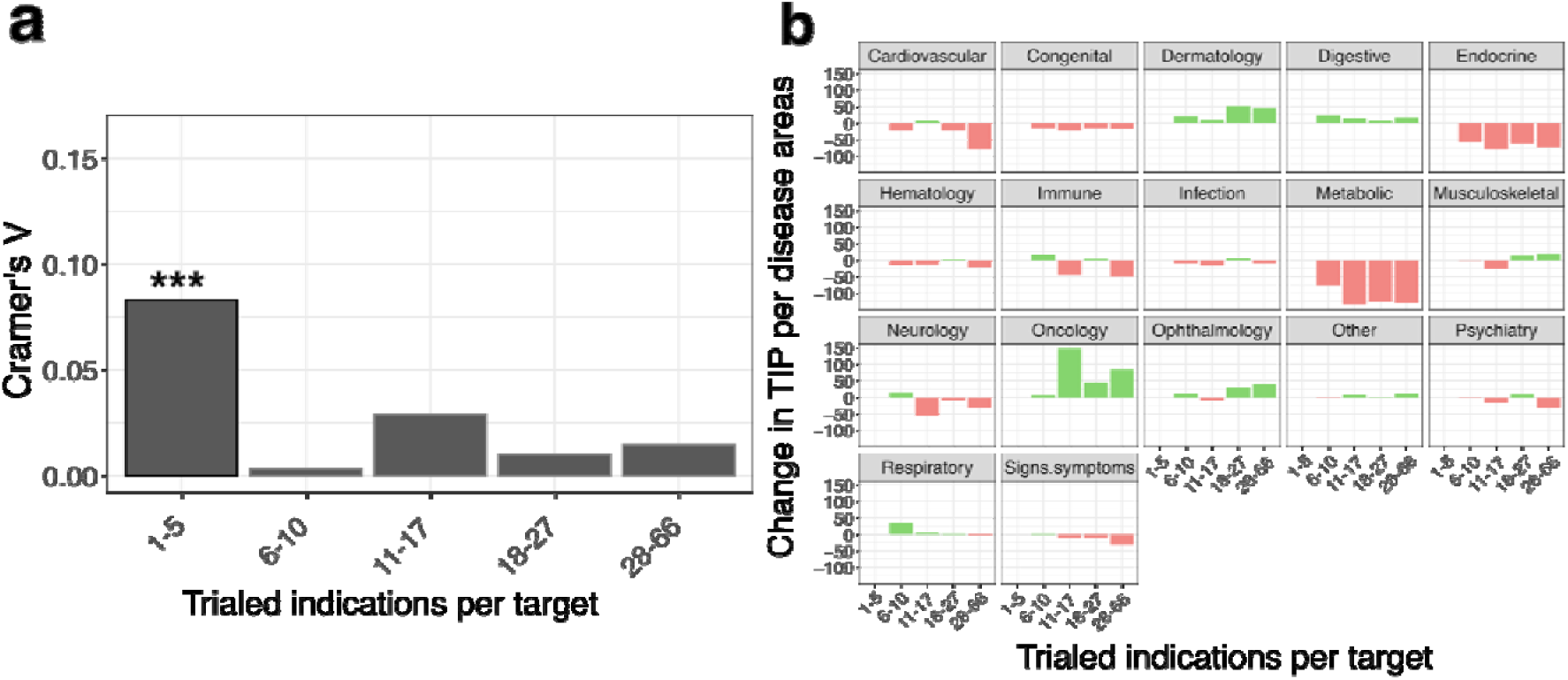
Target trial specificity differs between disease areas and affects MR predictivity. **(a)** Predictive power of the MR *P* value as given by Cramer’s V for targets grouped by the amount of indications they have been clinically trialed for (irrespective of trial outcome). Cramer’s V and *P* value are calculated based on the association between Phase II successful TIPs and significant MR results (ɑ = 0.0001). Negatives: Failed trials. Chi-squared test results indicated by asterisks. **(b)** Change in the number of TIPs associated with each disease area compared to the most specific target TIPs (1-5 tested indications). Bins have been chosen to be as equally sized as possible without overlaps. Bin sizes: n (1-5) = 2,337; n (6-10) = 2,318; n (11-17) = 2,281; n (18-27) = 2,311; n (28-66) = 2,235. Significancy asterisk code: * *P* < 0.05; ** *P* <0.01; *** *P* <0.001.

## References

1. Roland, A., Fox, W. & Baker, A. Efficiency, effectiveness and productivity in pharmaceutical R&D. Nat. Rev. Drug Discov. 23, 656–657 (2024).

2. Hay, M., Thomas, D. W., Craighead, J. L., Economides, C. & Rosenthal, J. Clinical development success rates for investigational drugs. Nat. Biotechnol. 32, 40–51 (2014).

3. Mullard, A. Parsing clinical success rates. Nat. Rev. Drug Discov. 15, 447 (2016).

4. Pearl, J. Causal inference in statistics: An overview. Stat. Surv. 3, 96–146 (2009).

5. Plenge, R. M., Scolnick, E. M. & Altshuler, D. Validating therapeutic targets through human genetics. Nat. Rev. Drug Discov. 12, 581–594 (2013).

6. Nelson, M. R. et al. The support of human genetic evidence for approved drug indications. Nat. Genet. 47, 856–860 (2015).

7. King, E. A., Davis, J. W. & Degner, J. F. Are drug targets with genetic support twice as likely to be approved? Revised estimates of the impact of genetic support for drug mechanisms on the probability of drug approval. PLoS Genet. 15, e1008489 (2019).

8. Minikel, E. V., Painter, J. L., Dong, C. C. & Nelson, M. R. Refining the impact of genetic evidence on clinical success. Nature 629, 624–629 (2024).

9. Ochoa, D. et al. Human genetics evidence supports two-thirds of the 2021 FDA-approved drugs. Nat. Rev. Drug Discov. 21, 551 (2022).

10. Angrist, J. D., Imbens, G. W. & Rubin, D. B. Identification of causal effects using instrumental variables. J. Am. Stat. Assoc. 91, 444–455 (1996).

11. Burgess, S., Butterworth, A. & Thompson, S. G. Mendelian randomization analysis with multiple genetic variants using summarized data: Mendelian randomization using summarized data. Genet Epidemiol 37, 658–665 (2013).

12. Burgess, S. et al. Using genetic association data to guide drug discovery and development: Review of methods and applications. Am. J. Hum. Genet. 110, 195–214 (2023).

13. Katan, M. B. Apolipoprotein E isoforms, serum cholesterol, and cancer. Lancet 1, 507–508 (1986).

14. Bowling, H., Cocucci, A., Koo, D. C. E. & Harrison, R. K. Analysis of phase II and phase III clinical trial terminations from 2013 to 2023. Nat. Rev. Drug Discov. (2025) doi:10.1038/d41573-025-00208-6.

15. Piñero, J., et al. DISGENET: Accelerating data-driven discovery in disease genomics and therapeutic development. bioRxiv 2026.01.05.697749 (2026) doi:10.64898/2026.01.05.697749.

16. Davies, N. M., Holmes, M. V. & Davey Smith, G. Reading Mendelian randomisation studies: a guide, glossary, and checklist for clinicians. BMJ 362, k601 (2018).

17. GTEx Consortium. The GTEx Consortium atlas of genetic regulatory effects across human tissues. Science 369, 1318–1330 (2020).

18. Yazar, S. et al. Single cell eQTL mapping identified cell type specific control of autoimmune disease. Science 376, (2022).

19. Murphy, A. E., Schilder, B. M. & Skene, N. G. MungeSumstats: a Bioconductor package for the standardization and quality control of many GWAS summary statistics. Bioinformatics 37, 4593–4596 (2021).

20. Schriml, L. M. et al. Human Disease Ontology 2018 update: classification, content and workflow expansion. Nucleic Acids Res. 47, D955–D962 (2019).

21. Fairley, S., Lowy-Gallego, E., Perry, E. & Flicek, P. The International Genome Sample Resource (IGSR) collection of open human genomic variation resources. Nucleic Acids Res. 48, D941–D947 (2020).

22. Interface to the OpenGWAS Database API. https://mrcieu.github.io/ieugwasr/.

23. Purcell, S. et al. PLINK: a tool set for whole-genome association and population-based linkage analyses. Am. J. Hum. Genet. 81, 559–575 (2007).

24. Bowden, J. et al. A framework for the investigation of pleiotropy in two-sample summary data Mendelian randomization. Stat. Med. 36, 1783–1802 (2017).

25. Pierce, B. L., Ahsan, H. & Vanderweele, T. J. Power and instrument strength requirements for Mendelian randomization studies using multiple genetic variants. Int. J. Epidemiol. 40, 740–752 (2011).

26. Burgess, S., Butterworth, A. & Thompson, S. G. Mendelian randomization analysis with multiple genetic variants using summarized data. Genet. Epidemiol. 37, 658–665 (2013).

27. Bowden, J., Davey Smith, G. & Burgess, S. Mendelian randomization with invalid instruments: effect estimation and bias detection through Egger regression. Int. J. Epidemiol. 44, 512–525 (2015).

28. Wald, A. The fitting of straight lines if both variables are subject to error. Ann. Math. Stat. 11, 284–300 (1940).

29. Agresti, A. An Introduction to Categorical Data Analysis_. (John Wiley & Sons, New York, 2007).

30. Wright, M. N. & Ziegler, A. Ranger: A fast implementation of random forests for high dimensional data in C++ and R. J. Stat. Softw. 77, (2017).

31. Chen, T. & Guestrin, C. XGBoost: A Scalable Tree Boosting System. arXiv [cs.LG] (2016) doi:10.48550/ARXIV.1603.02754.

